# Prescription drug monitoring programs increase racial/ethnic inequities in unmet demand for substance use disorder treatment among people who inject drugs. A repeated cross-sectional analysis of people who inject drugs in 19 US metro areas in 2012, 2015, 2018, and 2022

**DOI:** 10.1101/2025.06.10.25328800

**Authors:** Umedjon Ibragimov, Stephanie Beane, Regine Haardörfer, Danielle F. Haley, Courtney R. Yarbrough, Sabriya Linton, Leo Beletsky, Hannah L.F. Cooper

## Abstract

**Background:** Evidence indicates that prescription drug monitoring programs (PDMPs) reduce demand for substance use disorder (SUD) treatment among the general population, perhaps by minimizing the risk of SUD onset through limiting access to prescribed opioids. Little is known about PDMP effects on SUD treatment among people who inject drugs (PWID), a population at high overdose risk.

**Methods:** Using four waves (2012, 2015, 2018, and 2022) of National HIV Behavioral Surveillance (NHBS), we conducted two-way fixed-effect modelling of associations of state-level “mandated review” PDMP policies and individual-level (1) SUD treatment utilization, and (2) unmet demand for SUD treatment among 24,518 PWID in 13 states. We tested effect modification by race/ethnicity.

**Results:** PDMPs were associated with an 8 percentage-point increase in the probability of unmet demand for SUD treatment in the sample as a whole (95% CI: 3.0, 12.0). PDMP implementation was also associated with an increased Black, Indigenous, Latinx, and other people of color (BILPOC) vs. White gap in the probability of unmet demand, from a 3.0 percentage-point gap in non-PDMP states (95% CI: 1.0, 5.0) to a 9 percentage-point gap in PDMP states (95% CI: 7.0, 11.0).

**Conclusions:** PDMPs may increase racial/ethnic inequities in SUD treatment access. To strengthen PDMP effectiveness, supply reduction policies must be accompanied by enhanced access to SUD treatment and other services for PWID, particularly among BILPOC PWID.

## INTRODUCTION

Prescription drug monitoring programs (PDMPs) are among the most commonly enacted policies that state legislatures have deployed to curb the US overdose crisis, especially overdoses of prescription opioid pain relievers (PDMP TTAC, 2024; Rhodes et al., 2019). The age-adjusted rate of deaths from overdoses in the US increased from 8.9 per 100,000 standard population in 2003 to 32.6 in 2022 with slight decrease to 31.3 in 2023 (Garnett & Miniño, 2024). By 2022 every state had implemented a PDMP (PDMP TTAC, 2024). PDMPs are supply-side policies designed to curb the flow of controlled substances into communities and create state-sanctioned databases of individuals’ controlled substance prescribing and dispensing histories. At a minimum, states mandate that controlled substance dispensers (primarily pharmacists) collect and report prescription data to the PDMP. Many state legislatures have gone beyond this basic requirement and mandated prescribers (e.g., physicians) and dispensers (e.g., pharmacists) to check the patient’s individual record in the PDMP for prescription patterns associated with increased risk of overdose (e.g., concurrently prescribed opioids and benzodiazepines) or reflective of disordered use (e.g., multiple concurrent opioid prescriptions) before prescribing and dispensing controlled substances. This stipulation is known in the literature as “*mandated review*” or “*mandatory access*” (Grecu et al., 2019).

Multiple studies have examined the impact of PDMPs on substance use disorder (SUD) treatment participation as a proxy measure of SUD rates among the *general* population (Birk & Waddell, 2017; Grecu et al., 2019; Reifler et al., 2012; Reisman et al., 2009). To date, researchers have analyzed associations of these policies to population-level patterns of SUD treatment, and have found that “mandated review” PDMPs are associated with either overall reductions in population-level rates of SUD treatment admissions for prescribed opioids (Birk & Waddell, 2017; Grecu et al., 2019), or with slower increases in these rates (Reifler et al., 2012). These findings collectively suggest that PDMPs decrease SUD treatment enrollment in the general population by (1) reducing the diversion of prescription opioid analgesics and perhaps of other addictive substances, and thus (2) reducing the rate of SUD onset within the general population.

To our knowledge, no studies have analyzed the effects of PDMPs among people who inject drugs (PWID), a population at high risk of overdose (Colledge et al., 2019). Findings from PDMP research with general population samples may have limited generalizability to PWID. For example, while PDMPs may reduce the demand for SUD treatment among the general population by lowering the risk of SUD onset, they may ***increase*** the demand for SUD treatment among people who are already living with a SUD. Evidence suggests that limiting illegalized drug supplies may precipitate treatment seeking among PWID by increasing the risk of acute withdrawal (Mital et al., 2016; Zolopa et al., 2021). Because of limited access and barriers to SUD treatment in the US, including financial barriers, increased demand may be accompanied by increased *unmet* demand (Bouchery, 2017; Gertner et al., 2020; Han et al., 2017; Jones et al., 2015).

PDMPs may differentially increase both SUD treatment utilization and unmet demand for SUD treatment across White vs. Black, Indigenous, Latinx, and other people of color (BILPOC) PWID. PDMPs are more likely to curtail opioid supplies to BILPOC and thus may be more likely to precipitate withdrawal among BILPOC PWID. Biases and flaws in proprietary PDMP risk calculation algorithms disproportionately “red flag” BILPOC (Oliva, 2022). Townsend et al. (2022) found that PDMPs are associated with significant declines in high-dosage opioid analgesic prescription rates for Black patients, while White patients are unaffected, despite lower opioid analgesic prescription rates for Black patients prior to mandate implementation. However, one ecologic study reached contradictory results (Ayres & Jalal, 2018). Possibly, then, PDMPs create racial/ethnic differences in SUD treatment use and unmet demand among PWID

This analysis leverages four waves (2012, 2015, 2018, and 2022) of CDC’s National HIV Behavioral Surveillance (NHBS) to generate essential evidence about the relationships of “mandatory-access” PDMPs to (2) past-year SUD treatment use, and (2) unmet demand for SUD treatment among PWID (i.e., seeking but being unable to obtain treatment). The NHBS system allows us to extend beyond the current focus on general population samples, to assess the effect of PDMPs on individual PWID specifically. We also expand past PDMP analyses by assessing if PWID’s individual race/ethnicity moderates the relationship between PDMPs and SUD treatment outcomes.

## METHODS

### Overview, Units of Analysis, and Sample

We applied two-way, fixed-effect (state and year) models to explore associations between PDMP and SUD treatment and prescription drug misuse outcomes using serial cross-sectional data from NHBS and other existing administrative data. We have two units of analysis: (1) state, the geopolitical unit at which PDMP-mandated review laws are passed and implemented, and (2) individual PWID, the unit at which we measure outcomes and most covariates.

Data on individuals were drawn from four cycles (2012, 2015, 2018, and 2022) of NHBS PWID interviews. NHBS used respondent-driven sampling (RDS) to recruit approximately 500 PWID from each of 20 metropolitan statistical areas (MSAs) in 2012 and 2015, from each of 23 MSAs in 2018, and 19 MSAs in 2022. We used a two-year lag between PDMP policy adoption and NHBS data collection to account for the NHBS recall period and also for the full-scale implementation of PDMP policy. NHBS participant eligibility criteria for PWID include (1) injecting a non-prescribed drug in the past 12 months (NHBS does not query use of drugs prescribed to the individual); (2) being aged >18; (3) living in one of the participating MSAs; and (4) having capacity to complete the survey in English or Spanish. Additional information on NHBS eligibility and sampling is available elsewhere (Centers for Disease Control and Prevention, 2020; Lansky et al., 2007).

We limited our analytic sample to survey respondents in the 15 MSAs (in 13 states) where NHBS consistently sampled PWID in four interview cycles—2012, 2015, 2018, and 2022. Because NHBS does not query SUD diagnoses, we further limited the analytic sample to PWID who reported daily drug use, regardless of mode of administration (91.74% injected daily and 51.33% used non-injection drugs daily). We excluded PWID from the San Juan-Bayamon MSA because of their lack of racial/ethnic diversity and excluded participants with incomplete data on analytic variables (<3%; n=719).

### Measures

#### Outcomes

Both outcomes were drawn from dichotomous NHBS variables. One captured whether each participant reported engaging in SUD treatment in the past year (“Have you participated in a program to treat drug use in the past 12 months?”). The other captured past-year unmet demand for SUD treatment (“In the past 12 months, did you try to get into a program to treat drug use but were unable to?”).

#### Primary Independent Variable: Time-Varying State PDMP Mandated Prescriber Review

We measured PDMP mandated review requirements using a time-varying state-level dichotomous indicator, created using Prescription Drug Abuse Policy System (PDAPS) data (Prescription Drug Abuse Policy System, nd) based on the questions, “Does the state require prescribers to check the PDMP before prescribing controlled substances?” (2010-2014) or “Does the state require the PDMP to be queried for prescribing controlled substances or opioids/benzodiazepines” (2014-2019). PDMP data (and other drug-related policy data) in PDAPS was created using rigorous legal epidemiologic methods (e.g., legal analysts independently coding legal texts). We scanned the Council of the District of Columbia for PDMP mandated review legislation and identified March 2021 as the effective date (D.C. Law 23-251. Prescription Drug Monitoring Program Query and Omnibus Health Amendments Act of 2020). We lag the PDMP exposure measure from NHBS covariates by 24 months to account for (1) the year-long recall period for our outcome variables, and (2) an anticipated 12-month lag between adopting the PDMP mandated review and its hypothesized full-scale causal effect on the outcome.

#### Potential Effect Modifiers

We analyzed effect modification by individual-level race/ethnicity (White non-Hispanic/Latinx vs. BILPOC).

#### Individual-level Covariates

We operationalized the following individual-level covariates using NHBS data: sociodemographic characteristics (e.g., age, education, income), insurance status, history of incarceration (i.e., held in a detention center, jail, or prison >24 hours in past 12 months), unhoused status, documented NHBS HIV test result, duration of injection drug use, and employment (i.e., employed, unable to work due to health, not employed/other).

#### Time-Varying State-level Covariates

We included a dichotomous variable for state Medicaid expansion as of the 12-month NHBS lookback period for each NHBS wave (Kaiser Family Foundation, 2022).

### Analysis

We explored distributions of all variables and examined correlations. Age and number of years since first injection were highly correlated (*r*=0.80) so we eliminated age from models.

We analyzed the associations of the time-varying dichotomous indicator of state PDMP mandated review requirement (as of the 2-year lag described above) to 1) SUD treatment and 2) unmet demand for SUD treatment among PWID using a two-way (state and year) fixed-effect models for each outcome (Wooldridge, 2010). Including state and year fixed effects allows us to distinguish the possible impact of PDMPs from time-invariant confounding state heterogeneity and the common secular trend. We used linear probability models; we also tested models using a logit link for the dichotomous outcome and conclusions were unchanged. Due to the small number of states (n=13) in our analysis, we estimated confidence intervals and p-values with the wild cluster bootstrap method implemented using STATA’s boottest package (Roodman et al., 2019).

We analyzed the association between PDMPs and outcomes for the overall sample. We present unadjusted, adjusted, and adjusted models testing moderation for each outcome. We investigated whether the relationship between PDMP mandated review and each outcome varied by individual race/ethnicity using interaction terms. We probed significant (p < .05) interaction terms by estimating the probability of an outcome for all combinations of dichotomous PDMP variable and the relevant moderator along with group differences. Statistical analyses were carried out with STATA version 15.1.

### Ethics

Analyses were approved by the University’s Institutional Review Board (IRB).

## RESULTS

### Sample Description

The analytic sample resided in 13 states and included 24,518 PWID; there were 16,952 males and 7,379 females. Forty percent of participants were Black non-Hispanic/Latinx, 40% were White non-Hispanic/Latinx, 19% were Hispanic/Latinx, and < 2% were other races/ethnicities (Table 1). Most PWID experienced poverty: 78% reported a household income at or below the federal poverty level, and 65% were unhoused in the past 12 months.

**Table 1.** Participant characteristics, overall and by residence in a state and year with PDMP mandated review, for people who inject drugs (PWID) in 2012, 2015, 2018, and 2022, Centers for Disease Control and Prevention’s National HIV Behavioral Surveillance data (N=24,518^a^)

| **Characteristic** | **All**<br>**(n=24,518<sup>a</sup>)**<br>**N (%)** | **PDMP = No**<br>**(n=17,898)**<br>**N (%)** | **PDMP = Yes**<br>**(n=6,620)**<br>**N (%)** |
| --- | --- | --- | --- |
| **STATE (n=52 state-years)** |  |  |  |
| Medicaid expansion | 29 (56.77) | 15 (42.86) | 14 (82.36) |
| **INDIVIDUAL PWID (n=24,518)** |  |  |  |
| Received SUD<sup>b</sup> treatment | 9,777 (39.88) | 7,222 (40.35) | 2555 (38.60) |
| Unmet demand for SUD<sup>b</sup> treatment | 5,881 (23.99) | 4,109 (22.96) | 1772 (26.77) |
| Insured | 18,461 (75.3) | 12,905 (72.1) | 5556 (83.93) |
| Received new, sterile syringes from an SSP<sup>c</sup> | 14,197 (57.9) | 10,120 (56.54) | 4077 (61.59) |
| Age (yrs), mean (SD) | 44.74 (12.03) | 44.91 (12.27) | 44.27 (11.35) |
| Sex<sup>d</sup> |  |  |  |
| Male | 16,952 (69.14) | 12,487 (69.77) | 4465 (67.45) |
| Female | 7,379 (30.1) | 5,299 (29.61) | 2080 (31.42) |
| Missing | 187 (0.76) | 112 (0.63) | 75 (1.13) |
| Race/ethnicity<sup>d</sup> |  |  |  |
| Black non-Hispanic/Latinx | 9,777 (39.88) | 7,616 (42.55) | 2161 (32.64) |
| Hispanic/Latinx | 4,598 (18.75) | 3,050 (17.04) | 1548 (23.38) |
| White non-Hispanic/Latinx | 9,729 (39.68) | 6,925 (38.69) | 2804 (42.36) |
| Other | 414 (1.69) | 307 (1.72) | 107 (1.62) |
| Income at/below the federal poverty line | 19,008 (77.53) | 13,816 (77.19) | 5192 (78.43) |
| High-school graduate/GED | 17,080 (69.66) | 12,428 (69.44) | 4652 (70.27) |
| Employed full or part-time | 9,958 (40.62) | 7,578 (42.34) | 2,380 (35.95) |
| Incarceration (past 12 months) | 8,620 (35.16) | 6,518 (36.42) | 2102 (31.75) |
| Unhoused (past 12 months) | 15,906 (64.87) | 1,1205 (62.6) | 4701 (71.01) |
| No. years since 1st injection, mean (SD) | 21.05 (14.09) | 21.66 (14.36) | 19.41 (13.18) |
| Living with HIV | 1,678 (6.84) | 1,223 (6.83) | 455 (6.87) |

a. Overall sample reflects all four analysis years (2012, 2015, 2018, and 2022)
b. Substance use disorder
c. Syringe services program
d. Percentages may not add to 100% due to rounding

Of the 13 states in our sample, no states adopted a PDMP mandated review before June 2010 (the start of the exposure window for the 2012 NHBS sample). One state implemented mandatory PDMP review by June 2013 (start of the exposure window for the 2015 NHBS sample); 3 additional states implemented it by June 2016 (start of the exposure window for the 2018 NHBS sample); 8 additional states implemented it by June 2020 (start of the exposure window for the 2022 NHBS sample). Only D.C. had not adopted PDMP mandated review by the end of the exposure window for the study period (Supplemental Table 1).

Table 1 presents NHBS participant characteristics, comparing PWID in states and years with a PDMP mandated review to PWID in states in years without a mandate. Descriptive statistics indicate that the percentage of PWID reporting engaging in SUD treatment was similar in states and years with a mandated review (39%) compared to those without (40%) (Table 1). The percentage of PWID reporting unmet demand for drug treatment (27%) was higher in states and years with a mandated review compared to those without (23%).

### Model-based analysis

#### SUD treatment outcome

In the unadjusted models (Table 2), time-varying PDMP mandated review was not associated with past-year SUD treatment (Model 1: −5.0 percentage points; [95% CI, −14.0, 3.0]). The null relationship persisted in the adjusted model (Model 2: −4.0 percentage points; [95% CI, −13.0, 4.0]). The relationship between PDMP mandated review and SUD treatment enrollment did not vary by race/ethnicity (Model 3, Table 2).

**Table 2.** Model results for the estimated effect of residence in a PDMP mandated review state and other covariates on the probability of drug treatment, and unmet demand for drug treatment among people who inject drugs (PWID) in 2012, 2015, 2018, and 2022 Centers for Disease Control and Prevention’s National HIV Behavioral Surveillance (N=24,518)

|  | Probability of drug treatment<br>(95% CI<sup>b</sup>) *p* value | Probability of unmet demand for treatment<br>(95% CI<sup>b</sup>) *p* value |
| --- | --- | --- |
| **Model 1: Unadjusted model** |  |  |
| Residence in a PDMP state<sup>a</sup> | -0.05 (-0.14, 0.03), 0.20 | 0.05 (-0.001, 0.11), 0.05 |
| **Model 2: Adjusted<sup>c</sup> model** |  |  |
| Residence in a PDMP state<sup>a</sup> | -0.04 (-0.13, 0.04), 0.32 | 0.08 (0.03, 0.12), 0.002 |
| **Model 3: Adjusted<sup>c</sup> model including interaction with race/ethnicity** |  |  |
| Residence in a PDMP state<sup>a</sup> | -0.05 (-0.14, 0.03), 0.23 | 0.05 (0.002, 0.09), 0.04 |
| Race (ref: White non-Hispanic/Latinx) |  |  |
| BILPOC<sup>e</sup> | -0.009 (-0.05, 0.02), 0.55 | 0.03 (0.01, 0.05), 0.002 |
| PDMP state residence x race/ethnicity | 0.016 (-0.03, 0.07), 0.47 | 0.06 (0.04, 0.08), <0.001 |

a. Residence in a PDMP mandated review state (source PDAPS) as of the NHBS recall period for each year
b. The 95% confidence interval and *p*-values based on wild cluster bootstrapped models
c. Two-way fixed-effect (state and year) models adjusted for state Medicaid expansion and individual characteristics (receipt of clean needles from an SSP, insurance, gender, race/ethnicity, income, education, employment, incarceration, unhoused status, injection duration, HIV status, status as RDS seed, and network size)
d. Syringe service program
e. Black, Indigenous, Latinx and People of Color

#### Unmet demand for SUD treatment

In the unadjusted model (Table 2), PDMP mandated review was not associated with past-year unmet demand for SUD treatment at p < 0.05 (Model 1: 5.0 percentage points; [95% CI, −0.1, 11.0]). The relationship became statistically significant in the adjusted model (Model 2: 8.0 percentage points; [95% CI, 3.0, 12.0]).

In non-PDMP years, BILPOC PWID had a 3-percentage point greater probability of unmet demand compared to White non-Hispanic PWID (Table 3, Row B: 3.0 percentage points; [95% CI, 1.0, 5.0]). Post-PDMP the racial/ethnic difference increased and BILPOC PWID had a 9-percentage point greater probability of unmet demand than non-PDMP years (Table 3, Row A: 9.0 percentage points; [95% CI, 7.0, 11.0]). Comparing non-PDMP to PDMP state-years, there is an 11.0 percentage point increase in the probability of unmet demand for BILPOC PWID (Table 3, row C; [95% CI, 7.0, 15.0]) and a 5.0 percentage point increase for White PWID post-PDMP adoption (Table 3, row D [95% CI, 1.0, 8.0]).

**Table 3.** Interaction effects and differences: Probability of unmet demand for drug treatment by residence in a PDMP mandated review state and race/ethnicity among people who inject drugs (PWID) in the 2012, 2015, 2018, and 2022 Centers for Disease Control and Prevention’s National HIV Behavioral Surveillance (N=24,518)

|  | Probability of unmet demand for drug treatment (95% CI), P value |  |  |
| --- | --- | --- | --- |
|  | BILPOC | White non-Hispanic/Latinx | BILPOC – White difference |
| A. PDMP mandated review = Yes | 0.34 (0.30, 0.37), <0.001 | 0.25 (0.21, 0.28), <0.001 | 0.09 (0.07, 0.11), <0.001 |
| B. PDMP mandated review = No | 0.23 (0.21, 0.25), <0.001 | 0.19 (0.18, 0.22), <0.001 | 0.03 (0.01, 0.05), 0.001 |
|  | PDMP mandated review = Yes | PDMP mandated review = No | PDMP difference |
| C. BILPOC | 0.34 (0.30, 0.37), <0.001 | 0.23 (0.21, 0.25), <0.001 | 0.11 (0.07, 0.15), <0.001 |
| D. White non-Hispanic/Latinx | 0.25 (0.21, 0.28), <0.001 | 0.19 (0.18, 0.22), <0.001 | 0.05 (0.01, 0.08), 0.02 |

## DISCUSSION

Little is known about the effects of PDMP adoption on PWID, a population strongly affected by drug-related harms in the US, or about differential effects of PDMPs across racial/ethnic groups. By leveraging four cycles of the CDC’s NHBS data, we were able to test associations between PDMP mandated prescriber review adoption and unmet demand for and utilization of SUD treatment among PWID and examine sub-group differences in these associations. We found that adopting mandatory PDMP requirements was associated with increased unmet demand for SUD treatment among all PWID, with a greater impact among BILPOC PWID. Specifically, the unmet demand among White PWID increased by 5 percentage points, while BILPOC PWID experienced an 11-percentage point increase in unmet demand. At the same time, we found no association between PDMP adoption and SUD treatment utilization in the sample as a whole or within any single racial/ethnic group.

On the surface, it might seem contradictory that PDMPs simultaneously (1) did not change the probability of SUD treatment use among PWID, but (2) were associated with higher probabilities of ***unmet*** demand for SUD treatment. We offer the following interpretation for future empirical investigation. PDMPs limit the flow of controlled substances into communities (Moyo et al., 2017; Puac-Polanco et al., 2020). Curtailed flow could happen directly, to individual PWID who had been prescribed medications (Ali et al., 2017), but could also happen indirectly, via impacts on diverted medications into drug markets (Surratt et al., 2014). Past research on drug markets indicates that curtailed access to substances induces withdrawal among people with SUD (Mital et al., 2016; Zolopa et al., 2021). As a result, PDMP enactment may increase the demand for treatment among PWID. However, persistent barriers to SUD treatment may prevent PWID from successfully engaging in it. Well-documented barriers include limited spatial access to safety-net evidence-based treatment; stigma; and cost (Bouchery, 2017; Gertner et al., 2020; Han et al., 2017; Jones et al., 2015).

Our results indicate that BILPOC PWID experience greater increases in unmet demand for treatment than their White counterparts. Past literature suggests that PDMPs *disproportionately* diminish the flow of controlled substances into BILPOC communities (Townsend et al., 2022) through racially discriminatory implementation (Oliva, 2022). It is possible that BILPOC PWID who relied on this source, diverted or otherwise, would be more likely to experience gaps in access to controlled substances and thus to experience drug withdrawal than their White counterparts. Curtailed supply may lead to increased *interest* in engaging in SUD treatment among BILPOC PWID. These PWID, however, may not be able to *access* care to the same extent as their White counterparts because of the US’s racialized SUD treatment system (Stahler et al., 2021; Substance Abuse and Mental Health Services Administration. Office of Behavioral Health Equity, 2020). Multiple processes undergird this racialization. Health insurance inequities make SUD treatment unaffordable for many BILPOC patients. Even when individuals are insured, residential segregation and lack of providers in predominantly BILPOC areas erect spatial barriers to accessing SUD treatment (Cook & Alegría, 2011; Cooper et al., 2016; Goedel et al., 2020; Weinstein et al., 2017). Collectively, racialized PDMP implementation coupled with racialized access to SUD treatment may explain why PDMPs disproportionately increase unmet demand for SUD treatment among BILPOC PWID.

The strengths of our study include a repeated cross-sectional design, which allowed us to capture changes in the underlying demographic composition of PWID in these metro areas over time. The multilevel nature of the study allowed us to analyze covariates at the state, ZIP, and individual levels. The study featured a large racially/ethnically diverse sample of PWID recruited in 19 metropolitan areas, which allowed us to analyze the impact of a state law to individual outcomes in this vulnerable population and associated inequitable effects. The main limitation of the repeated cross-sectional design is inability to assess the effect of PDMP adoption on within-person *changes* in the outcomes. Moreover, findings may have little generalizability to PWID residing in rural areas or in states not included in our sample. Self-reported data may be prone to measurement error due to recall and social desirability biases; however, we assume that it is unlikely that this measurement error would be of systematic nature and differ based on the state’s PDMP policies.

## Conclusion

PDMPs are a popular policy instrument adopted in many states that are designed to help curb the US overdose epidemics. Our findings suggest that PDMPs may increase unmet demand for SUD treatment among PWID, with greater adverse impacts among BILPOC PWID. These findings add to a large body of literature suggesting that supply reduction policies should be implemented in tandem with efforts to enhance access to SUD treatment and other harm reduction services, with a particular focus on addressing racialized, systemic barriers to access to care.

## Data Availability

NHBS data are held at the CDC, and individuals must request access
via that agency.

