## Supplemental Table 1 for "Prescription drug monitoring programs increase racial/ethnic inequities in unmet demand for substance use disorder treatment among people who inject drugs. A repeated cross-sectional analysis of people who inject drugs in 19 US metro areas in 2012, 2015, 2018, and 2022"

| **Supplemental Table 1: Years of PDMP prescriber mandated review timing, 2012, 2015, 2018, and 2022 Centers for Disease Control and Prevention’s National HIV Behavioral Surveillance data (N=15 states)** | |
| --- | --- |
| **State^a^** | **Year of PDMP mandated review** |
| California | 2017 |
| Colorado | 2018 |
| District of Columbia | 2021^b^ |
| Georgia | 2018 |
| Illinois | 2018 |
| Louisiana | 2014 |
| Maryland | 2018 |
| Michigan | 2018 |
| New Jersey | 2015 |
| New York | 2013 |
| Pennsylvania | 2015 |
| Texas | 2015 |
| Washington | 2018 |
| a. Sample includes 15 MSAs (Atlanta, GA; Baltimore, MD; Chicago, IL; Houston, TX; Denver, CO; Detroit, MI; New York City, NY; New Orleans, LA; Newark, NJ; Philadelphia, PA; Los Angeles, San Diego, & San Francisco, CA; Seattle, WA; Washington, DC) from 13 states that participated in all four (2012, 2015, 2018, 2022) NHBS waves.  b. Did not meet the PDMP implementation cut off for NHBS 2022 (June 2020 cut off) | |
